# Methylomic, proteomic, and metabolomic correlates of traffic-related air pollution: A systematic review, pathway analysis, and network analysis relating traffic-related air pollution to subclinical and clinical cardiorespiratory outcomes

**DOI:** 10.1101/2023.09.30.23296386

**Authors:** Cameron Casella, Frances Kiles, Catherine Urquhart, Dominique S. Michaud, Kipruto Kirwa, Laura Corlin

## Abstract

A growing body of literature has attempted to characterize how traffic-related air pollution (TRAP) affects molecular and subclinical biological processes in ways that could lead to cardiorespiratory disease. To provide a streamlined synthesis of what is known about the multiple mechanisms through which TRAP could lead cardiorespiratory pathology, we conducted a systematic review of the epidemiological literature relating TRAP exposure to methylomic, proteomic, and metabolomic biomarkers in adult populations. Using the 139 papers that met our inclusion criteria, we identified the omic biomarkers significantly associated with short-or long-term TRAP and used these biomarkers to conduct pathway and network analyses. We considered the evidence for TRAP-related associations with biological pathways involving lipid metabolism, cellular energy production, amino acid metabolism, inflammation and immunity, coagulation, endothelial function, and oxidative stress. Our analysis suggests that an integrated multi-omics approach may provide critical new insights into the ways TRAP could lead to adverse clinical outcomes. We advocate for efforts to build a more unified approach for characterizing the dynamic and complex biological processes linking TRAP exposure and subclinical and clinical disease, and highlight contemporary challenges and opportunities associated with such efforts.

## 1. Introduction

It is well-established that exposure to traffic-related air pollution (TRAP) is associated with adverse respiratory and cardiovascular outcomes [1–3]. Research suggests that the pathways underlying associations between TRAP exposure and cardiorespiratory outcomes likely involve oxidative stress, endothelial dysfunction, and inflammatory responses [4–10]. A growing number of epidemiological studies are investigating how (changes in) DNA methylation patterns (methylomics), proteomic profiles, and metabolomic profiles underlie the physiological pathways linking TRAP exposure to respiratory and cardiovascular health (e.g., [11–16]). Nevertheless, no large-scale longitudinal study to date has identified common biological pathways involving TRAP-related methylomic, proteomic, and metabolomic patterns - evidence that could help establish a unified multi-omics framework to gain a better understanding of the adverse health consequences of air pollutants and designing relevant interventions.

Previous work has outlined many of the challenges of establishing a unified multi-omics approach to air pollution epidemiology (e.g., need for repeated samples, identification of an appropriate exposure metric, availability of appropriate statistical techniques to handle the large number of omics analytes) [17–21]. Additionally, when comparing and synthesizing results from across studies, challenges related to heterogeneity in study designs, populations, air pollutants of interest, exposure windows, omics measurement methods, and analytic techniques arise [11,12,21–24]. Despite these challenges, multi-omics integration (i.e., integrating across multiple levels of biology such as methylation patterns, proteomic profiles, and metabolomic profiles) aimed at understanding mechanisms linking environmental risk factors to chronic disease can advance clinical and public health knowledge and inform design and implementation of relevant interventions [25–27]. Therefore, as a step towards the larger goal of developing an integrated multi-omics approach, we conducted the first systematic review of studies relating TRAP to three types of omics signals. Using these signals from across omics types, we aimed to pinpoint common biological pathways known to be involved in respiratory and cardiovascular diseases, assess challenges and benefits of a multi-omics approach, and identify research needs. The number of studies directly linking TRAP exposure to clinical outcomes through changes in omics signals is relatively small; however, we believe that identifying omics signals and pathways known to be associated with both TRAP exposure and cardiorespiratory disease is a prudent step towards advancing clinical and public health decision making.

## 2. Materials and Methods

### Search Strategy and Study Selection

We searched Embase and PubMed for English-language epidemiologic articles published between January 2010 and February 2023 that reported on the association between TRAP exposure and one or more of three omics types (DNA methylation [methylomics], proteomics, and metabolomics). Search terms included DNA methylation, proteomics, metabolomics, TRAP, and particulate matter (PM). The search strategy is described in detail in Supplementary S1. We screened extracted articles by title and abstract. We excluded reviews and reports, as well as in-vitro, in-silico, ex-vivo, and animal studies. We excluded articles not containing one or more TRAP exposures, and those that examined “pollution” without specifying pollutants. Relevant pollutants included particulate matter < 2.5 microns (PM_2.5_), particulate matter < 10 microns (PM_10_), PM constituents, ultrafine particulate matter (UFP), black carbon (BC), elemental carbon (EC), organic carbon (OC), nitrogen dioxide (NO_2_), nitrogen oxides (NOx), carbon monoxide (CO), sulfur dioxide (SO_2_), sulfate (SO_4_^2-^), ozone (O_3_), diesel exhaust (DE), and polycyclic aromatic hydrocarbons (PAHs). Studies focused on people who were pregnant or under 18 years of age were also excluded. In addition to 115 articles that remained after screening, we identified 24 papers through expert knowledge, for a total of 139 unique studies. There were 54 methylomic, 57 proteomic, 37 metabolomic, and 9 overlapping studies—four of which included both proteomics and metabolomics, and five that included both proteomics and methylation (Figure 1).

**Figure 1:**
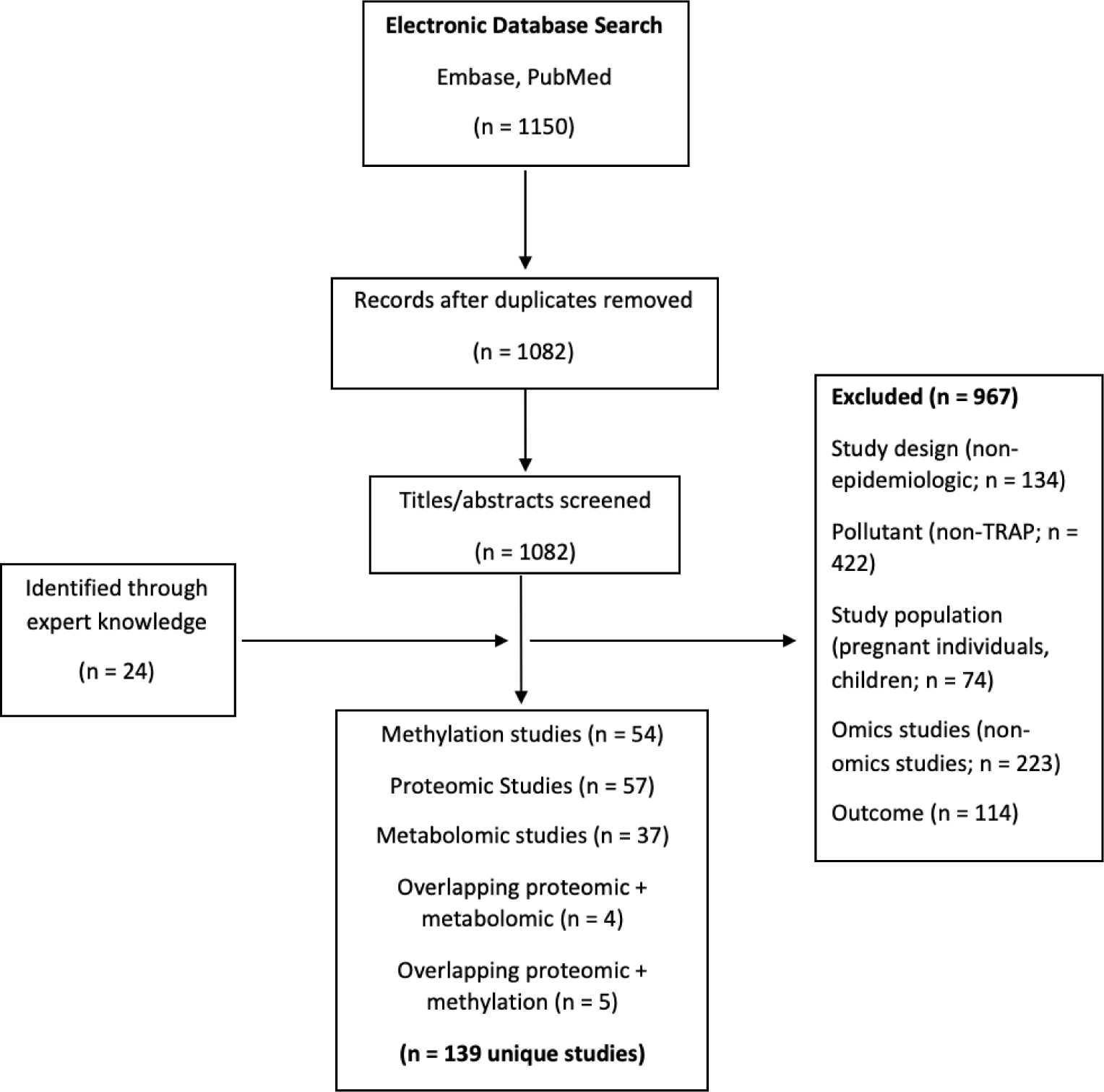
Flow diagram of the article selection process with exclusion criteria.

### Data Extraction and Organization

We extracted the following from each article: study design and sample size, air pollution exposure methods, exposure metrics, omics assay methods, participant demographics, statistical methods, and results (Table 1 and Supplementary S2 Tables 1-3). Statistically significant associations between different TRAP exposures and each omics article type (methylomic, proteomic, metabolomic) were identified (Supplementary S2, Tables 4-6). We used statistical significance thresholds determined by the original authors, which included both adjusted and non-adjusted p-values. Air pollution exposures were split by pollutant type and averaging period (short-term: ≤30 days; long-term: >30 days).

**Table 1:** Overview of the literature.

| Omics Type | Study Design | Exposure Assessment | Exposure Window | Study Populations<sup>a</sup> | Country | Sample Size | Sex Distribution | Omics Approach |
| --- | --- | --- | --- | --- | --- | --- | --- | --- |
| Methylomics<br>n = 54 studies | Cross-sectional: 29<br>Panel: 9<br>Cohort: 5<br>Cross-over: 9<br>Quasi-experimental: 2 | Fixed site measurement: 16<br>Spatiotemporal model: 21<br>Personal measurement: 12<br>Controlled exposure: 5 | Short-term: 29<br>Long-term: 25 | NAS: 10 [38–47]<br>KORA: 3 [42,46,48]<br>WHI: 3 [49–51]<br>ARIC: 3 [49–51]<br>EPIC-Italy: 2 [52,53]<br>MESA: 2 [54,55]<br>Sister Study: 2 [56,57]<br>BAPE: 2 [58,59]<br>Taiwan Biobank: 2 [60,61]<br>REGICOR: 1 [52]<br>EPIC-Netherlands: 1 [53]<br>Lifelines: 1 [48]<br>EXPOsOMIC S: 1 [62]<br>SAPALDIA: 1 [63]<br>Lothian Birth Cohort: 1 [64]<br>SPHERE: 1 [65] | USA: 17<br>China: 15<br>Italy: 8<br>Canada: 4<br>Netherlands: 3<br>Taiwan: 3<br>Germany: 2<br>Switzerland: 2<br>UK: 2<br>Belgium: 2<br>Spain: 1<br>South Korea: 1<br>Czech Republic: 1 | <50: 20<br>50-99: 3<br>100-1000: 20<br>>1000: 11 | 100% female: 4<br>100% male: 11<br>Other: 39 | Candidate gene: 26<br>Epigenome-wide association study: 24<br>Global methylation: 4 |
| Proteomics<br>n = 57 studies | Cross-sectional: 28<br>Panel: 8<br>Cohort: 3<br>Cross-over: 10<br>Quasi-experimental: Case-control: 3 | Fixed site measurement: 24<br>Spatiotemporal model: 19<br>Personal measurement: 9<br>Biomarker: 2<br>Controlled exposure: 4 | Short-term: 36<br>Long-term: 21 | NAS: 3 [66–68]<br>SWAN: 3 [69–71]<br>KORA: 3 [72–74]<br>Heinz-Nixdorf Recall: 3 [72,75,76]<br>Framingham Offspring: 2 [77,78]<br>AIRCHD: 2 [79,80]<br>EPIC-Italy: 1 | USA: 17<br>China: 17<br>Canada: 6<br>Germany: 4<br>India: 3<br>Taiwan: 3<br>Italy: 2<br>Sweden: 1<br>UK: 1<br>France: 1<br>Brazil: 1<br>Sweden: 1<br>Finland: 1<br>Switzerland: 1 | <50: 15<br>50-99: 10<br>100-1000: 13<br>>1000: 19 | 100% female: 3<br>100% male: 6<br>Other: 48 | Targeted: 54<br>Untargeted: 3 |
|  |  |  |  | [81]<br>BPRHS: 1<br>[82]<br>Malmo Diet<br>and Cancer: 1<br>[83]<br>AHAB-II: 1<br>[84]<br>SAGE: 1 [85]<br>Nurse's<br>Health Study:<br>1 [86]<br>ELISABET: 1<br>[87]<br>ESCAPE: 1<br>[88]<br>SAPALDIA: 1<br>[72]<br>FINRISK: 1<br>[72]<br>TwinGene: 1<br>[72]<br>MESA: 1 [89]<br>CAFEH: 1 [90]<br>CoLaus: 1<br>[91] |  |  |  |  |
| Metabolomics<br>N = 37 studies | Cross-sectional: 15<br>Panel: 7<br>Cohort: 2<br>Cross-over: 7<br>Natural Experiment: 1 | Fixed site<br>measurement: 8<br>Spatiotemporal model:<br>10<br>Personal<br>measurement: 14<br>Biomarker: 1<br>Controlled exposure: 4 | Short-term: 26<br>Long-term: 11 | DRIVE: 3 [92–<br>94]<br>NAS: 2<br>[95,96]<br>Children's<br>Health Study:<br>2 [97,98]<br>KORA: 2<br>[99,100]<br>SAPALDIA: 1<br>[101]<br>EPIC-Italy: 1<br>[101]<br>ACE: 1 [102]<br>ACE-2: 1<br>[103]<br>Oxford St. 2: 1<br>[104]<br>TAPAS II: 1<br>[104]<br>CAFEH: 1<br>[105]<br>EARTH: 1<br>[106]<br>AIRCHD: 1<br>[80]<br>SCOPE: 1<br>[107]<br>TwinsUK: 1<br>[108] | USA: 17<br>China: 12<br>Germany: 2<br>UK: 2<br>Sweden: 1<br>Switzerland: 1<br>Italy: 1<br>India: 1<br>Spain: 1<br>Netherlands: 1<br>Brazil: 1 | <50: 15<br>50-99: 6<br>100-1000: 7<br>>1000: 4 | 100% female: 1<br>100% male: 5<br>Other: 31 | Targeted: 8<br>Untargeted: 29 |

<sup>a</sup>Numbers represent the number of papers reviewed containing the given characteristic. Where the original study included multiple study populations, all study populations and countries were counted. Abbreviations: ACE – Atlanta Commuters Exposure; AHAB-II – Adult Health and Behavior; AIRCHD – Air Pollution and Cardiovascular Dysfunctions in Healthy Adults Living in Beijing; ARIC – Atherosclerosis Risk in Communities; BPRHS – Boston Puerto Rican Health Study; CAFEH – Community Assessment of Freeway Exposure and Health; DRIVE – Dorm Room Inhalation to Vehicle Emissions; EARTH – Environmental and Reproductive Health; ELISABET - Enquête Littoral Souffle Air Biologie Environnement; EPIC – European Prospective Investigation into Cancer and Nutrition; ESCAPE – European Study of Cohorts for Air Pollution Effects; KORA – Cooperative Health Research in the Region of Augsburg; MESA – Multiethnic Study of Atherosclerosis; NAS – Normative Aging Study; REGICOR - REGistre Glroní del COR; SAGE – Study on Global Aging and Adult Health; SAPALDIA - Swiss Study on Air Pollution and Lung Disease in Adults; SCOPE - A Prospective Study COMparing the Cardiometabolic and Respiratory Effects of Air Pollution Exposure on Healthy and Prediabetic Individuals; SPHERE - Susceptibility to Particle Health Effects, miRNA and Exosomes; SWAN – Study of Women's Health Across the Nation; TAPAS – Transportation, Air Pollution and Physical Activities; WHI – Women's Health Initiative

Using the significant associations shown in Supplementary S2 Tables 4-6, we identified common biological processes and type of biomarker represented across the omics types (abbreviated version of results shown in Table 2 and full results shown in Supplementary S2 Table 7). Gene Ontology (GO) molecular functions (molecular level activities performed by gene products, e.g., glucose transmembrane transport) were extracted for each gene and protein [28]. Where available, Kyoto Encyclopedia of Genes and Genomes (KEGG) pathways (pathways of common molecular interaction, e.g., tumor necrosis factor signaling) were indicated for all genes, proteins, and metabolites [29–31]. For genes and proteins without KEGG data, GO biological processes (functions of gene products) were used instead. The nextprot knowledgebase [32] was used to extract GO molecular functions, GO biological processes, and KEGG pathways for all genes and proteins. The GenomeNet KEGG COMPOUND Database [33] was used to extract KEGG functions for all available metabolite markers. Based on Supplementary S2 Tables 4-7, we created a simplified conceptual diagram of the putative relationships among TRAP, omics signals, subclinical processes, and clinical outcomes (Figure 2).

**Table 2:** Combined synthesis of significant associations. Abbreviations: Al – Aluminum, BC – Black Carbon, CO – Carbon Monoxide, CO_2_ – Carbon Dioxide, Cu – Copper, DE – Diesel Exhaust, Fe – Iron, K – Potassium, Ni – Nickel, NO – Nitrogen monoxide, NO_2_ – Nitrogen dioxide, NO_x_ – Nitric oxides, O_3_ - Ozone, Pb – Lead, PM – Particulate matter, Se – Selenium, Si – Silicon, SO_4_ - Sulfate, TRAP – Traffic-related air pollution, UFP – Ultrafine particulate matter, V – Vanadium, Zn – Zinc.

**Figure 2:**
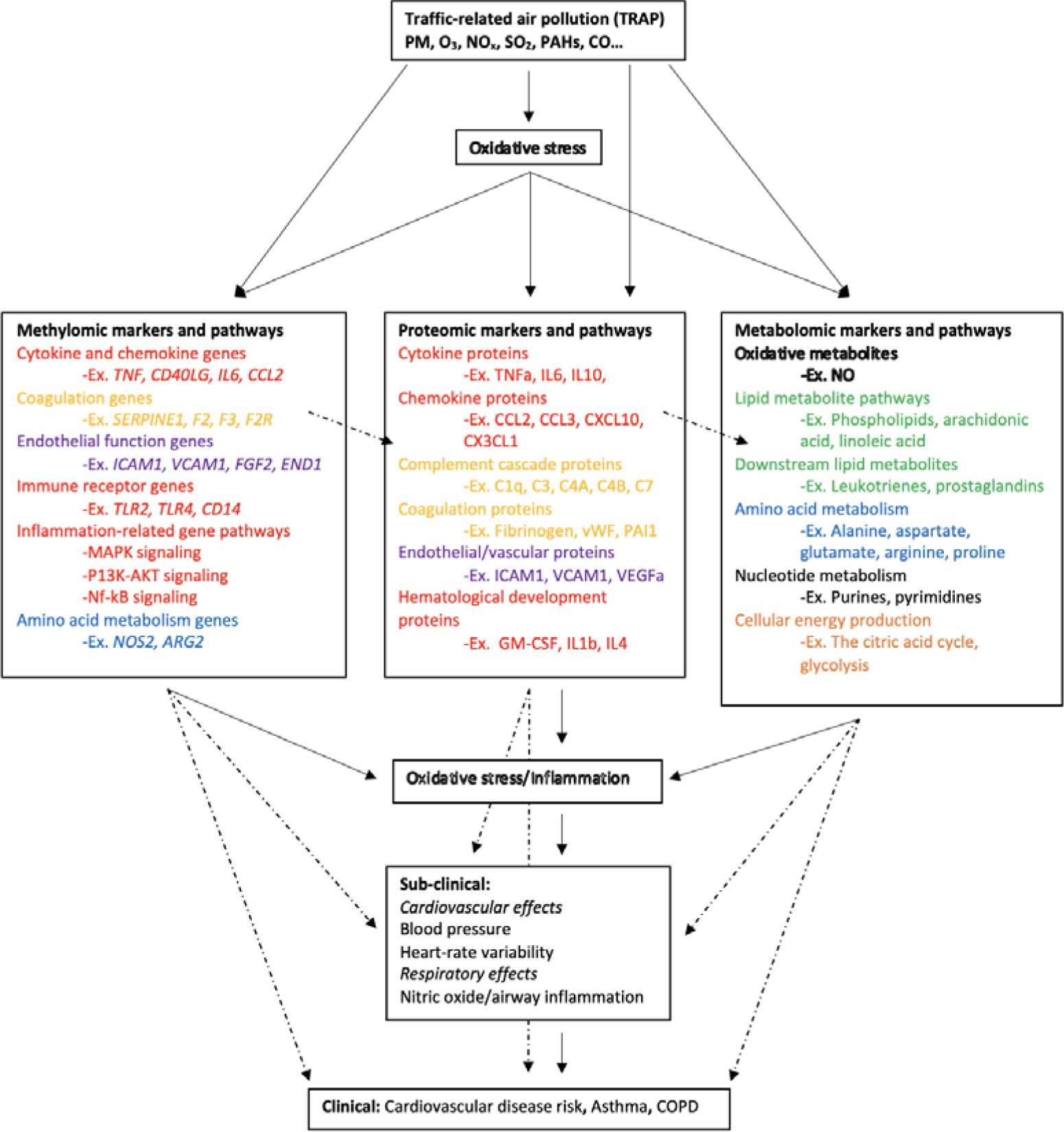
Overview of the relationships among traffic-related air pollution, omics markers, and subclinical and clinical cardiovascular and respiratory disease outcomes. Solid arrows indicate a well-established, known relationship as evidenced by the biomedical literature. Dashed arrows indicate a probable association or an association with possible mediators that needs to be further investigated. Color coding of text within methylomic, proteomic, and metabolomic text boxes corresponds to a category of biological pathway. Green - lipid metabolism; orange - cellular energy production; blue - amino acid metabolism; red – inflammation and immunity; yellow - coagulation, purple - endothelial function; white - oxidative stress; black - analytes that do not clearly fit into the above categories (vitamins, purines, xanthines, etc.) Abbreviations: ARG2 – Arginase 2; C1q – Complement component 1q; C3 – Complement component 3; C4A - Complement component 4A; CCL2 – CC motif chemokine ligand 2/monocyte chemoattractant protein 1; CCL3 – CC motif chemokine ligand 3/macrophage inflammatory protein 1 alpha; CD14 – Cluster of differentiation 14; CD40LG – Cluster of differentiation 40 ligand; CX3CL1 – Fractalkine; CXCL10; CXC motif chemokine ligand 10/interferon gamma inducible protein 10; F2 – Coagulation factor 2; F2R-Coagulation factor 2 receptor; F3 – Coagulation factor 3; FGF2 – Fibroblast growth factor 2; GM-CSF – Granulocyte macrophage colony stimulating factor; ICAM1-Intercellular adhesion molecule 1; IL1b – Interleukin 1 beta; IL4 – Interleukin 4; IL6 – Interleukin 6; IL10 – Interleukin 10; MAPK – Mitogen activated protein kinase; NOS2 – Nitric oxide synthase 2; Nf-KB – Nuclear factor kappa light chain enhancer of activated B cells; P13K-AKT – Phosphatidylinositol 3 kinase and AKT/protein kinase B; SERPINE1 – Serpin family E member 1/Plasminogen activator inhibitor 1; TLR2 – Toll like receptor 2; TLR4 – Toll like receptor 4; TNF-Tumor necrosis factor alpha; TNFa – Tumor necrosis factor alpha; VCAM1 – Vascular cell adhesion molecule 1; VEGFa – Vascular endothelial growth factor alpha; vWF – Von Willebrand factor.

### Pathway and Network Analyses

We conducted bioinformatics analyses synthesizing the results across the omics studies using the lists of relevant biomarkers shown in Supplementary S2 Table 8 (representing all significant associations shown in Supplementary S2 Tables 4-6). We used the open-source tools Reactome [34] and MetaboAnalyst 5.0 [35] to conduct pathway analyses. Specifically, we used Reactome to perform overrepresentation pathway analyses on the gene methylation sites and proteins that were significantly associated with TRAP exposure (separately for each omic type and for associations with short- and long-term TRAP exposures), and MetaboAnalyst to conduct a KEGG pathway analysis of all metabolites that were significantly associated with TRAP (separately for short- and long-term exposures). These programs generate lists of pathways indicated by the extracted analytes. Some pathways discussed in this review were not on the indicated lists of these pathway analyses, and therefore statistical significance values were not given. Given that we extracted the KEGG functions and/or GO data for each analyte, we were able to group together omics signals effectively, despite pathway analysis-related statistical thresholds that may be limiting in representing all biological pathways involved in TRAP-exposure.

MetaboAnalyst was also used to conduct four KEGG network analyses representing the functional relationships among biomarkers. We created two networks incorporating methylation markers and metabolites that were significantly associated with short- and long-term TRAP exposure and two networks incorporating proteins and metabolites that were significantly associated with short- and long-term TRAP exposure (in each case, using separate networks for short- and long-term exposures). In network analyses, networks are parameterized by degree (i.e., number of incoming/outgoing edges on each node) and betweenness (i.e., number of shortest paths between each pair of nodes). Higher values for degree and betweenness restrict the network to only the most highly connected and relevant nodes [36,37]. For our two short-term network analyses, degree and betweenness filters were constrained to degree of at least three. In the long-term exposure analyses, networks did not contain enough nodes to apply these filters. This is due to the relative sparsity of literature examining associations between long-term exposures and omics signals.

## 3. Results and Discussion

### 3.1 Overview of the Literature

Table 1 provides an overview of the study designs, exposure assessment approaches, study populations, sample sizes, sex distributions, and omics approaches used in the studies included in this review.

#### 3.1.1 TRAP Exposure Assessment

Exposure assessment approaches differed by omics type: spatiotemporal modeling was most common for methylomic papers, fixed site monitoring was most common for proteomics papers, and personal monitoring was most common for metabolomics papers (Table 1). Short-term exposures were more commonly assessed than long-term exposures for each omic type. For long-term exposures, the most common exposure window was an annual average. As in air pollution epidemiology generally, each exposure assessment approach and exposure window has strengths and weaknesses in the context of different study designs; a potential benefit of a multi-omics approach is the enhanced reliability of knowledge obtained from triangulating findings from studies that employ the diverse combinations of exposure assessment techniques and windows.

The most common pollutant studied across all three omics (regardless of exposure window) was PM_2.5_. Forty-six methylation papers, 41 proteomics papers, and 32 metabolomics papers measured PM_2.5_ exposure. PM_10_, UFP, BC, NO_2_, NO_x_, and O_3_ were all considered in each omic type; however, they were less commonly studied in papers focused on long-term exposures. Papers that did not investigate PM_2.5_ generally focused on O_3_ or diesel exhaust. Given the study designs and exposure assessment methods, time-varying exposures and TRAP mixtures were generally not accounted for in the analyses; future studies should consider time-varying exposures and mixtures.

#### 3.1.2 Study Populations

Research in this field predominantly draws from populations in North America, China, and Western Europe (Table 1); future studies should include more geographic diversity – requiring an investment in TRAP exposure and omics assessment in other geographic regions. Additionally, although most study populations included people regardless of sex, single-sex cohorts were common (especially for methylomic papers where 28% were single-sex). Three methylomic, two proteomic, and four metabolomic papers considered effect modification by sex [42,54,64,83,98,109–112] (Supplementary S2 Tables 1-3). Fourteen methylomic, 16 proteomic, and 21 metabolomic studies contained populations with a mean age or entire age range of 35 years old or younger. Twenty-three methylomic, nine proteomic, and four metabolomic studies contained populations with a mean age, or entire age range of 60 years or older. In general, the methylomic literature had slightly older participants and metabolomics literature had slightly younger participants. However, there was adequate representation of all ages throughout all three omics types. Most studies included healthy participants or did not specify health conditions as criteria for eligibility.

#### 3.1.3 Biological Matrices

Methylomic, proteomic, and metabolomic markers were assessed using a variety of biological matrices, (Supplementary S2 Tables 1-3). Leukocytes and whole blood were the most common biological matrices for methylomic papers (27 and 17 papers, respectively). All studies adjusted for cell composition except those exclusively using CD4+ helper cells or buccal cells as the matrix of interest, or those using paired samples with a short lag time [113–118]. For proteomic papers, serum and plasma were the most common biological matrices (34 and 21 papers, respectively). Nine proteomics papers used both serum and plasma, with the inclusion of plasma serving primarily to measure fibrinogen levels [69,72,78,89,119–123]. Three proteomics papers used bronchoalveolar lavage fluid to understand the associations between TRAP and the bronchoalveolar proteome, serving as a more direct measure of TRAP’s influence [124–126]. Similar to proteomics, serum and plasma were the most common biological matrices for metabolomics papers (17 and 14 papers, respectively). Serum is currently considered the gold standard in metabolomics research, providing more sensitive results in biomarker detection; however, plasma also provides accurate results and has high reproducibility [127,128]. Five metabolomics papers utilized urine [98,129–132] and two used bronchoalveolar lavage fluid [133,134].

In general, decisions about the biological matrix were largely determined based on availability of data within a cohort rather than on the biological relevance of a given matrix for TRAP-cardiorespiratory relationships. Although other matrices (e.g., myocytes, bronchiolar cells, endothelial cells, etc.) may serve as a more direct source of omics signals, they are often inaccessible and/or invasive to procure [135,136]. Additionally, none of the studies explicitly considered biomarker interactions (e.g., protein-protein or protein-metabolite) or the possibility of biomarker degradation or metabolism (e.g., considering how TRAP exposure may only affect biomarker levels over a specific temporal window) [135,137–139]. Finally, without the ability to obtain repeated measures of multiple omics types within individuals over relevant time periods, it is not possible to directly assess putative relationships between TRAP exposure and cascading biological processes. That is, although we can view the associations among multiple omics layers and pollutants across similar short- and long-term exposure windows, we do not have a direct means to measure the exact temporal changes in methylomic, proteomic, and metabolomic makers occurring at consistent points post-exposure.

#### 3.1.4 Omics Assessment

In the methylomics literature, multiple high-throughput approaches and bioinformatics technologies were used (Supplementary S2 Table 1). The most common forms of methylation quantification were methylation arrays (37 papers) and bisulfite polymerase chain reaction (PCR) sequencing (13 papers). The PCR sequencing papers focused on candidate gene approaches (primarily for inflammatory and immune-related proteins, as well as genes related to circadian rhythm and epigenetic age) [38,47,116,117,120,140–147]. Analyses using arrays took advantage of the evolving technology to capture the most comprehensive set of biomarkers possible: one paper utilized a 385K array [43], 24 utilized a 450K array [39–42,44–46,48,50–55,57,62–64,148–153], and 12 utilized an 850K array [58–61,114,115,154–159]. Although we recommend the use of the most comprehensive technology available, the contribution of groundbreaking studies using older arrays to the current body of knowledge should not be understated [160,161]. Similarly, for the bioinformatics analyses of the methylomics results, researchers took advantage of the rapidly evolving tools such as KEGG for pathway analysis [39,43,114,133,157,158], the National Institutes of Health Databases for Annotation, Visualization and Integrated Discovery (NIH-DAVID) [39,53,62,152,153], Ingenuity Pathway Analysis (IPA) [40,63,126,148,154,162], Mummichog [15,92–94,101–103,105,106,163,164], and MetaboAnalyst [95–97,112,130,165–168].

Compared to the methylomics literature, there was homogeneity in approaches used across the proteomics literature (Supplementary S2 Table 2). Only three of the 57 proteomics papers used untargeted omics approaches (and therefore, the use of bioinformatics approaches for analysis were limited to relatively few studies) [126,132,162]. Instead, many studies assessed the concentration of approximately 20 targeted proteins (e.g., cytokines, chemokines, and other immune/inflammatory-related markers). This led to abundant data on the associations among TRAP and the concentration of key proteins related to inflammation and immunity, and therefore cardiorespiratory disease. The represented proteins often overlapped well with the proteins encoded by candidate genes targeted in methylation studies. While this is useful for multi-omics interpretation, the relative lack of untargeted analyses may limit our understanding of the complete proteomic response to TRAP, and potentially bias our analyses by over-representing certain processes already considered important in cardiorespiratory disease. Furthermore, it can make it difficult to integrate methylomic, proteomic, and metabolomic results together.

In contrast to the proteomics literature, most (28/37) of the metabolomics papers used untargeted approaches and 22 incorporated bioinformatics approaches for the interpretation of results (e.g., 11 used Mummichog [15,92–94,101–103,105,106,163,164] and nine used MetaboAnalyst [95–97,112,130,165–168]; Supplementary S2 Table 3). Specific to metabolomics is the challenge of metabolite identification. Fourteen of 37 metabolomics papers had level one confidence (highest level of confidence confirmed by reference standard) [80,94,97,100–102,106–108,110,134,163,168,169], whereas an additional six studies contained some level one matches mixed with lower confidence findings [93,103–105,164,170]. Thirteen studies had level two confidence, primarily confirmed by library spectrum match [15,96,112,118,129,130,133,165–167,171–173]. Only two studies did not contain metabolites with level two or greater confidence [96,98]. The variation in metabolite identification confidence reflects a level of uncertainty in the metabolomics signals observed across different studies [174,175].

### 3.2 Omics Markers and Associated Biological Pathways

Omics markers representing biological pathways related to lipid metabolism, cellular energy production, amino acid metabolism, inflammation and immunity, coagulation, endothelial function, and oxidative stress were present across the literature. In this section, we outline trends in common biological pathways and molecular functions associated with methylomic, proteomic, and metabolomic markers of TRAP exposure, along with the hypothesized connections to cardiorespiratory disease. Not all omics markers may be related to clinical outcomes, and further research is needed to identify the most critical pathways underlying the relationship between TRAP exposure and disease. Figure 2 shows a simplified diagram of the relationships. The supporting literature is summarized in Table 2 and Supplementary S2 Tables 4-7.

Table 2 synthesizes the methylomic, proteomic, and metabolomic literature together. The table is organized by KEGG pathway, and only includes those pathways most represented in the literature: lipid metabolism, cellular energy production, amino acid metabolism, inflammation and immunity, coagulation, endothelial function, and oxidative stress. Within each KEGG pathway, all methylomic, proteomic, and metabolic markers significantly associated with short- and/or long-term TRAP are noted. Each omics type was separated into associations for short- and long-term exposure. Details are given in the following sections.

#### 3.2.1 Lipid Metabolism

Phospholipids, sphingolipids, and acylcarnitines were represented throughout the metabolomics literature. However, no studies explored the associations between TRAP and methylomic or proteomic markers related to lipid metabolism (Table 2; Supplementary 2 Tables 6-7). In the metabolomics literature, both short- and long-term PM_2.5_ exposures were negatively associated with phospholipid levels [26–30]. In contrast, short-term UFP, NO_2_, and O_3_ were consistently and positively associated with levels of phospholipids [95,100,176]. Phospholipid metabolism is essential for normal cellular function as it is involved in generating biological membranes and plays an important role in cellular signaling processing in nearly all tissues [177]. Phospholipid imbalances are implicated in neurological disorders and neurodegenerative diseases, while damaged and oxidized phospholipids are associated with atherosclerosis and cardiovascular disease (CVD; Figure 2) [178,179]. It is not understood exactly how TRAP associations with phospholipid metabolites contribute to the aforementioned diseases.

Sphingolipids, such as sphingosines and some sphingomyelins, were negatively associated with short- and long-term PM_2.5_ as well as with short-term UFP [95,98,168], but were positively associated with short-term O_3_ and Ni [95,98,176,180]. For example, sphingosine 1-phosphate (a known risk factor for coronary artery disease (CAD)) [181] was negatively associated with short-term UFP and positively associated with short-term Ni [95]. Additionally, ceramide (a reaction product of sphingomyelin and/or sphingosine that is elevated in patients with hypertension, angina pectoris, myocardial infarction, and stroke [182–184]) was negatively associated with short-term PM_2.5_ and UFP exposure [95,168]. However, eight sphingomyelins were positively associated with long-term PM_2.5_ and short-term O_3_ [95,176]. Given these findings, it is possible that TRAP (and particularly the PM components) may not predominately work through pathways involving sphingolipids to affect CVD. However, future studies should confirm this hypothesis and should also consider whether methylation patterns or proteins related to lipid metabolism are implicated.

In contrast to the trends with sphingolipids, acylcarnitines were positively associated with short-term TRAP and negatively associated with short-term NO_2_ [95,104,111,132,165,170,173]. It has been shown that higher levels of medium and long-chain acylcarnitines are positively associated with both CVD, and risk of cardiovascular death in patients with stable angina pectoris [185–187].

Although most markers of lipid metabolism were considered only in the metabolomics literature, arachidonic acid and linoleic acid metabolism KEGG pathways were considered in both the proteomics (one protein involved in each) and metabolomics (20 and 13 metabolites, respectively) literature (Table 2). Synthesizing the results from these studies, our MetaboAnalyst pathway analyses suggested that the arachidonic acid metabolism KEGG pathway was significantly enriched by metabolites associated with both short- and long-term TRAP exposure (p = 4.29 × 10^−4^ and p = 0.01, respectively). Specifically, exposure to short-term diesel exhaust was associated with higher concentrations of the protein arachidonate 15-lipoxygenase (ALOX15). This enzyme helps generate bioactive lipid molecules, such as eicosanoids, hepoxilins, and lipoxins [188]. Interestingly, short-term diesel exhaust was also associated with lower levels of multiple metabolites related to ALOX15 [126,134]. The metabolomics literature also considered other components of the arachidonic acid and linoleic acid metabolism pathways. For example, short-term PM_2.5_ and diesel exhaust exposure were associated with higher and lower levels of eicosanoids, respectively [107,134]. These signaling lipids regulate homeostatic and inflammatory processes, making them important markers in the progression of CVD [188,189]. Additionally, short-term PM_2.5_ and other TRAP exposures were associated with higher levels of thromboxane, prostaglandin, and leukotriene metabolites [98,134,164,165,169].

These metabolites are associated with modification of the immune and inflammatory responses, and help mediate leukocyte accumulation [190]. Finally, short-term PM_2.5_, NO_2_, and other short-term TRAP exposures, as well as long-term PM_2.5_ and NO_2_ were associated with higher levels of metabolites involved in linoleic acid metabolism [99,100,134,164,165,167]. Dysregulated linoleic acid metabolism is traditionally considered pro-inflammatory and pathological, but the linoleic acid pathway is still not well understood [189].

The network analyses we conducted consistently identified metabolites related to arachidonic and linoleic metabolism, such as arachidonic acid, ALOX15, leukotrienes, prostaglandins, and thromboxanes (Figures 3-6; green symbols correspond to lipid metabolism). These metabolites associated with short-term air pollution exposures were connected with genes and proteins related to inflammation and the immune system (red symbols), endothelial function (pink symbols), and coagulation (yellow symbols; Figures 3 and 5). Lipid metabolism markers associated with long-term air pollution exposures had similar trends, though fewer nodes were identified for the gene-metabolite network overall (Figures 4 and 6).

**Figure 3:**
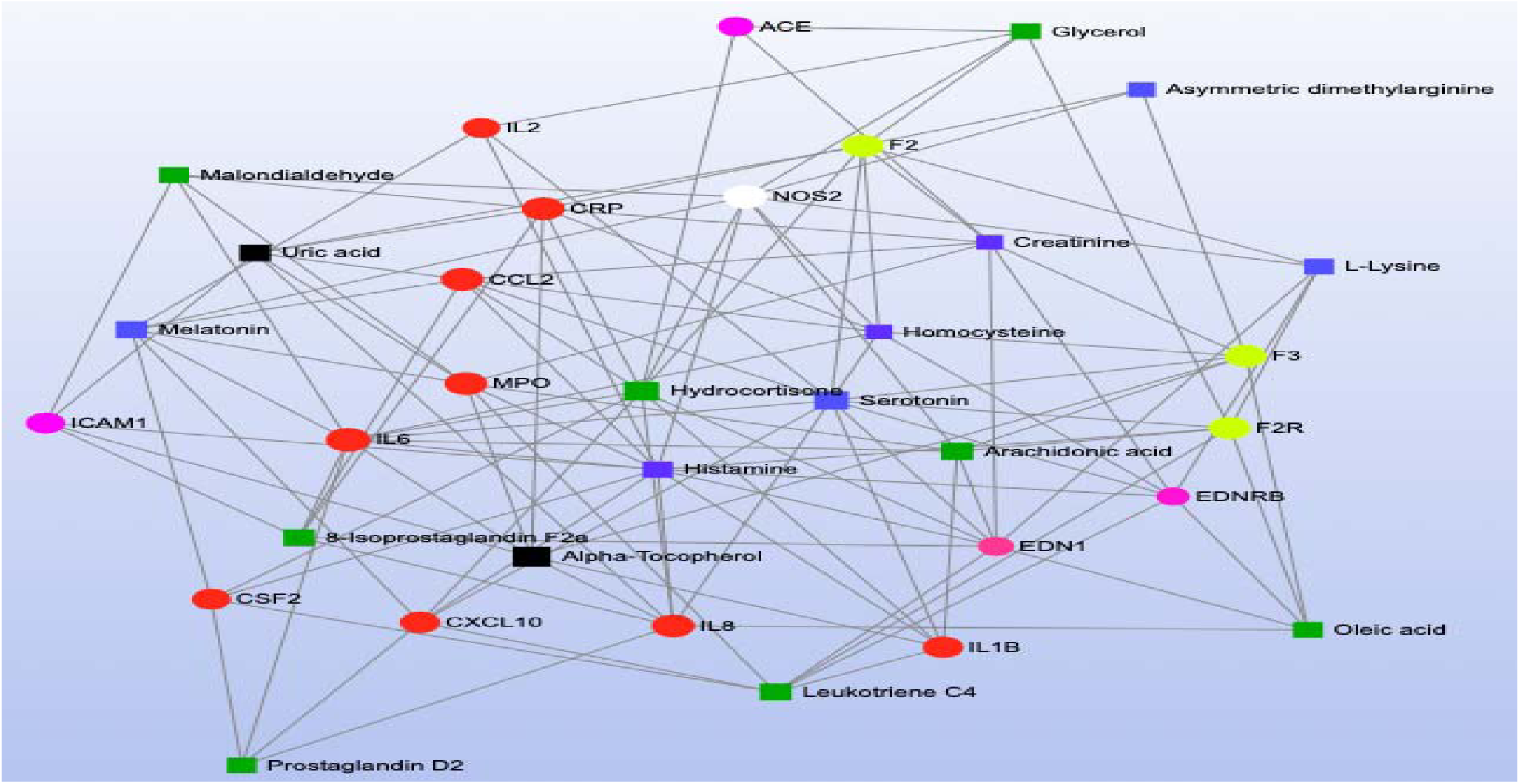
Short-term air pollution and gene-metabolite network analysis. Circular nodes represent genes, whereas square nodes represent metabolites. The color of each node corresponds to the category of biological pathway to which that analyte belongs. Green - lipid metabolism; orange - cellular energy production; blue - amino acid metabolism; red – inflammation and immunity; yellow - coagulation, pink - endothelial function; white - oxidative stress; black - analytes that do not clearly fit into the above categories (vitamins, purines, xanthines, etc.). Abbreviations: ACE – Angiotensin converting enzyme; CCL2 – Monocyte chemoattractant protein 1; CRP – C-reactive protein; CSF2 – Colony stimulating factor 2; CXCL10 – Interferon gamma induced protein 10; EDN1 – Endothelin 1; EDNRB – Endothelin receptor type B; F2 – Coagulation factor 2; F2R-Coagulation factor 2 receptor; F3 – Coagulation factor 3; IL1B – Interleukin 1 beta; IL −2 Interleukin 2; IL6 – Interleukin 6; IL-8; Interleukin 8; ICAM1 – Intercellular adhesion molecule 1; MPO – Myeloperoxidase; NOS2 – Nitric oxide synthase 2.

**Figure 4:**
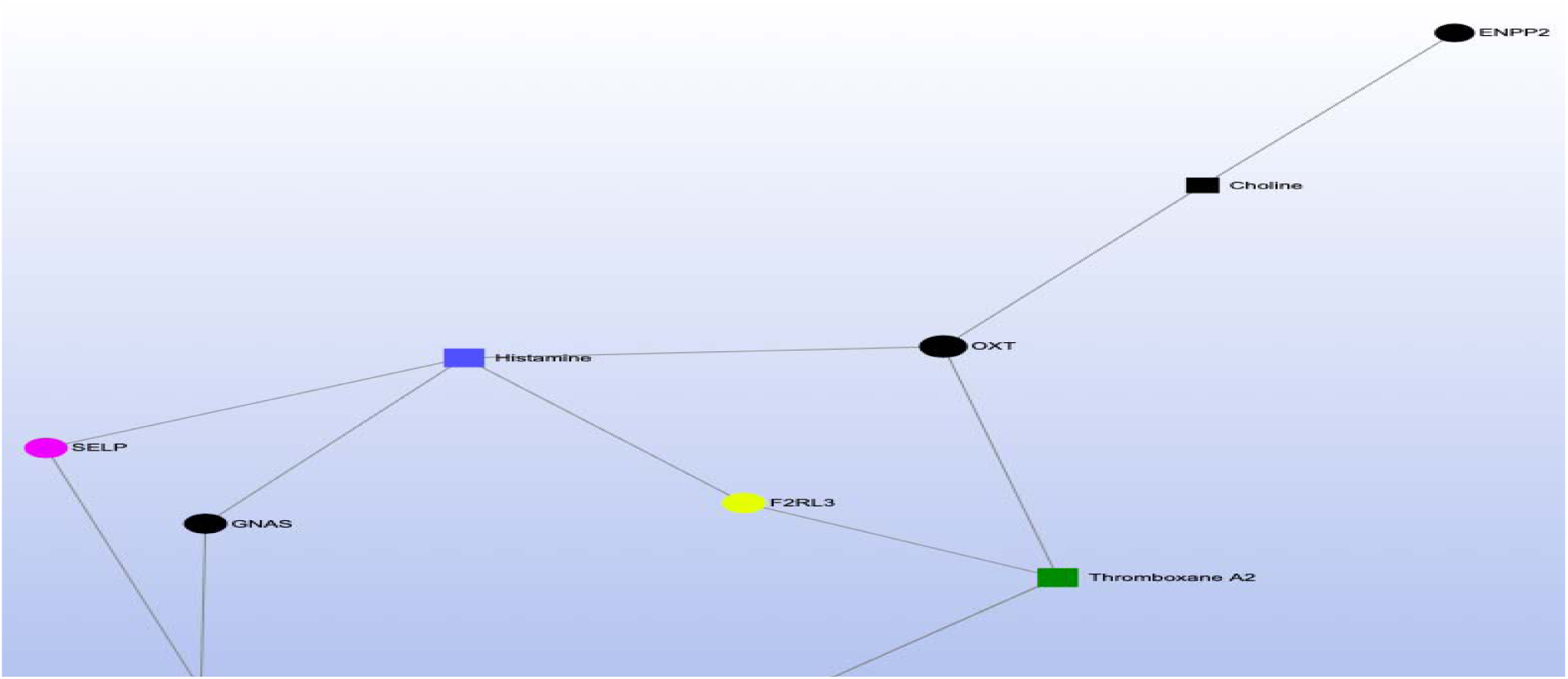
Long-term air pollution and gene-metabolite network analysis. Circular nodes represent genes, whereas square nodes represent metabolites. The color of each node corresponds to the category of biological pathway to which that analyte belongs. Green - lipid metabolism; orange - cellular energy production; blue - amino acid metabolism; red – inflammation and immunity; yellow - coagulation, pink - endothelial function; white - oxidative stress; black - analytes that do not clearly fit into the above categories (vitamins, purines, xanthines, etc.). Abbreviations: CACNA2D1 – Calcium voltage gated channel auxiliary subunit alpha2delta 1; ENPP2 – Ectonucleotide pyrophosphatase 2; F2RL3 – Coagulation factor 2 receptor like thrombin or trypsin receptor 3; GNAS – GNAS complex locus; OXT – Oxytocin prepropeptide; SELP – P selectin.

**Figure 5:**
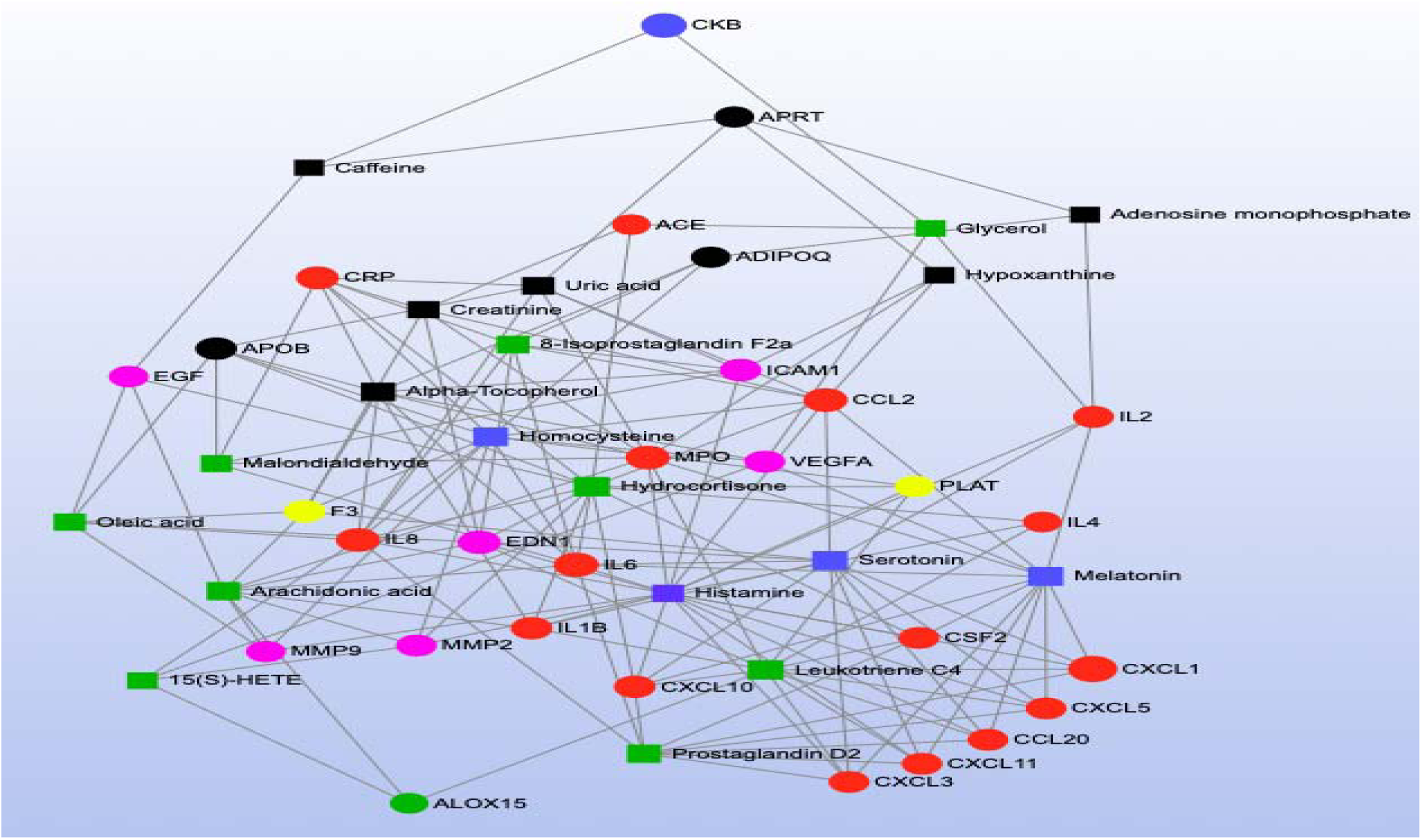
Short-term air pollution and protein-metabolite network analysis. Circular nodes represent proteins, whereas square nodes represent metabolites. The color of each node corresponds to the category of biological pathway to which that analyte belongs. Green - lipid metabolism; orange - cellular energy production; blue - amino acid metabolism; red – inflammation and immunity; yellow - coagulation, pink - endothelial function; white - oxidative stress; black - analytes that do not clearly fit into the above categories (vitamins, purines, xanthines, etc.). Abbreviations: 15(3)-HETE – 15 Hydroxyeicosatetraenoic acid; ACE – Angiotensin converting enzyme; ALOX15 – Arachidonate 15 lipoxygenase; APRT – Adenine phosphoribosyltransferase; APOB – Apolipoprotein B; CCL2 – monocyte chemoattractant protein 1; CCL20 – CC motif chemokine ligand 20; CKB – Creatine kinase B; CRP – C reactive protein; CSF2 – Colony stimulating factor 2; CXCL1 – CXC motif chemokine ligand 1; CXCL3 – CXC motif chemokine ligand 3; CXCL5 – CXC motif chemokine ligand 5; CXCL10- Interferon gamma induced protein 10; CXCL11 – CXC motif chemokine ligand 11; EGF-Epidermal growth factor; EDN1 – Endothelin 1; F3 - Coagulation factor 3; IL1B – Interleukin 1 beta; IL2 – Interleukin 2; IL4 – Interleukin 4; IL6 – Interleukin 6; IL8 – Interleukin 8, ICAM1 – Intercellular adhesion molecule 1; MMP2 – Matrix metalloproteinase 2; MMP9 – Matrix metalloproteinase 9; MPO – Myeloperoxidase; PLAT – Plasminogen activator, tissue type; VEGFA – Vascular endothelial growth factor A.

**Figure 6:**
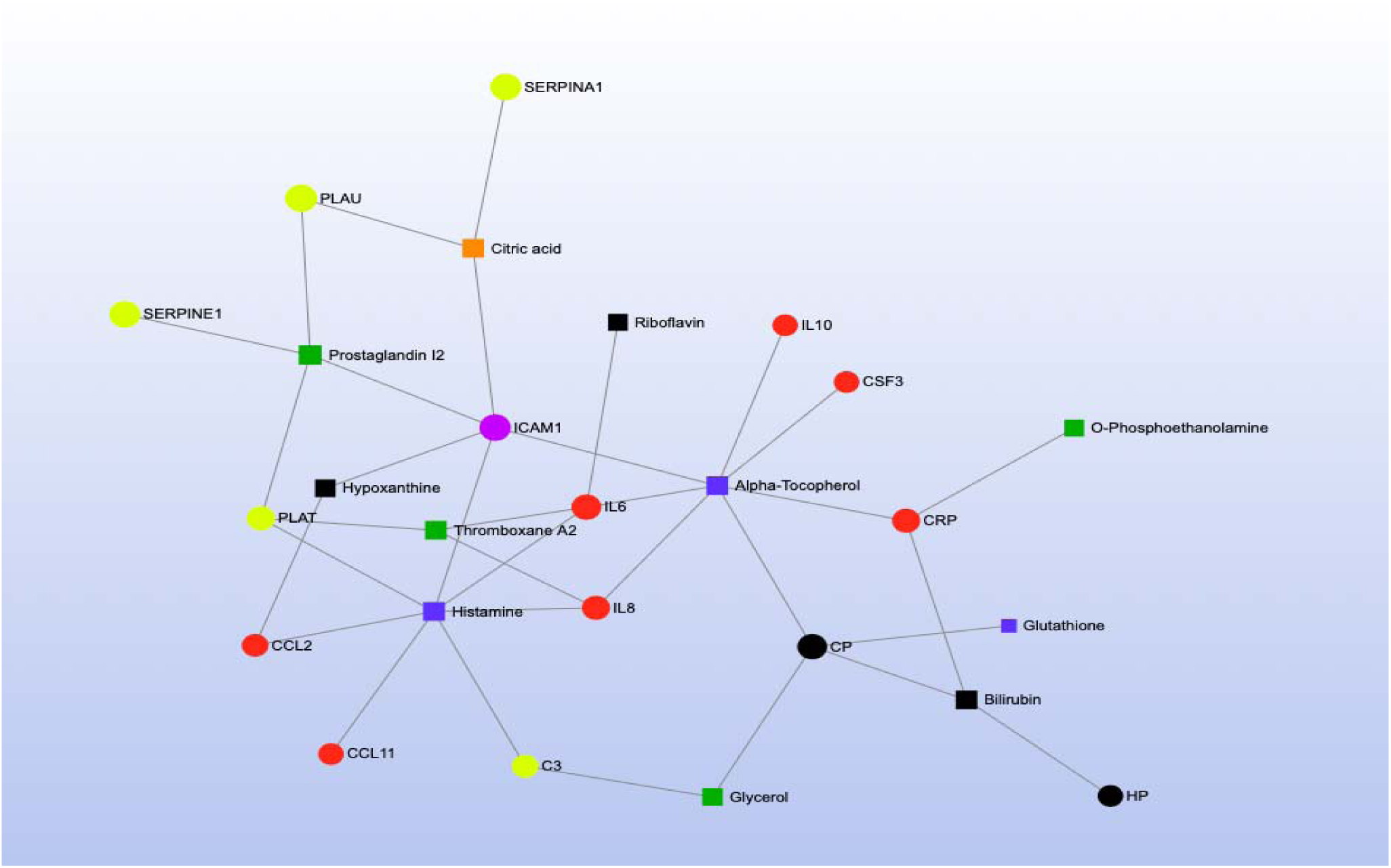
Long-term air pollution and protein-metabolite network analysis. Circular nodes represent proteins, whereas square nodes represent metabolites. The color of each node corresponds to the category of biological pathway to which that analyte belongs. Green - lipid metabolism; orange - cellular energy production; blue - amino acid metabolism; red – inflammation and immunity; yellow - coagulation, pink - endothelial function; white - oxidative stress; black - analytes that do not clearly fit into the above categories (vitamins, purines, xanthines, etc.). Abbreviations: C3 – Complement component 3; CCL2 – Monocyte chemoattractant protein 1; CCL11 – CC motif chemokine ligand 11; CP – Ceruloplasmin; CRP-C reactive protein; CSF3 – Colony stimulating factor 3; HP – Haptoglobin; ICAM1 – Intercellular adhesion molecule 1; IL6 – Interleukin 6; IL8 -Interleukin 8; IL10 – Interleukin 10; PLAT - Plasminogen activator, tissue type; PLAU – Plasminogen activator, urokinase; SERPINA1 – Alpha 1 proteinase inhibitor; SERPINE1 – Plasminogen activator inhibitor 1.

#### 3.2.2 Cellular Energy Production

Three cellular energy production KEGG pathways were associated with short- and long-term TRAP exposure: (1) the citric acid cycle, (2) glycolysis/gluconeogenesis, and (3) the pentose phosphate pathway (Table 2, Figure 2). Although no methylomic or proteomic markers related to the citrate cycle were identified as significantly associated with TRAP, our MetaboAnalyst pathway analyses synthesizing results across studies identified the citric acid cycle KEGG pathway as being significantly enriched by the metabolites significantly associated with short- and long-term TRAP exposure (p = 8.86 × 10^−3^ and p = 1.65 × 10^−3^, respectively). Specifically, exposure to short-term TRAP was associated with higher levels of some citric acid cycle intermediates (e.g., succinyl-CoA, succinate, cis-aconitic acid, and alpha-ketoglutaric acid) [130–132,191] but short-term PM_2.5_ exposure was associated with lower levels of pyruvate, while short-term EC was associated with lower levels of citric acid and isocitric acid [94]. In contrast, long-term PM_2.5_ exposure was associated with higher levels of malic acid and succinic acid [95,163]. Notably, citric acid cycle dysregulation has been associated with CVD [192,193]. For example, one case-cohort study found an increased risk of CVD with higher concentrations of fasting plasma malic acid, 2-hydroxyglutarate, and fumarate [193], while a nested case-control study found higher levels of succinic acid, malic acid, citric acid, and 2- hydroxyglutarate to be associated with a higher risk of atrial fibrillation [192]. Higher levels of malic acid and succinic acid associated with long-term PM_2.5_ exposure may underlie part of the known association between TRAP and the risk of CVD. Future studies could explore whether TRAP exposure is also associated with the methylation of genes encoding for key rate limiting and regulatory enzymes in the citric acid cycle, such as citrate synthase, isocitrate dehydrogenase, and alpha-ketoglutarate dehydrogenase, as well the concentrations of these enzymes. Additionally, future studies could explore functional relationships among citric acid and coagulation and endothelial function given the relationships we identified in the long-term air pollution and protein-metabolite network analysis (Figure 6).

The central carbohydrate metabolism pathways represented by biomarkers associated with TRAP include the glycolysis/gluconeogenesis and pentose phosphate pathways (Figure 2). The glycolysis/gluconeogenesis KEGG pathway was represented by two proteomic and five metabolomic markers significantly associated with TRAP, but no methylomic markers (Table 2). Similarly, five metabolomic (but no methylomic or proteomic markers) identified as belonging to the pentose phosphate KEGG pathway were significantly associated with TRAP (Table 2). For the glycolysis/gluconeogenesis KEGG pathway, exposure to short-term diesel exhaust was associated with lower levels of the protein alcohol dehydrogenase class four mu/sigma chain, and higher levels of the protein aldehyde dehydrogenase dimeric nicotinamide adenine dinucleotide phosphate-preferring [126]. In metabolomics studies, exposure to short-term PM_2.5_ was associated with lower levels of the metabolites lactate, pyruvate, and glyceric acid 1,3-bisphosphate [94,129,194], and exposure to long-term PM_2.5_ was associated with lower levels of 3-phosphoglycerate and lactate [95]. Short-term exposure to O_3_ was associated with higher levels of glucose and lactate [176], whereas exposure to short-term TRAP was associated with lower levels of glucose and 3-phosphoglycerate [95,132]. For the pentose phosphate KEGG pathway, short-term PM_2.5_, PM components, and certain other TRAP exposures were associated with lower levels of the metabolites glyceraldehyde, glycerate, 3- phosphoglycerate, and pyruvate [94,95,194], and long-term PM_2.5_ was associated with lower levels of glycerate and 3-phosphoglycerate [94,95,108,132,163,176,194]. However, short-term exposure to O_3_ was associated with higher levels of glucose and glycerate [176]. In pathological circumstances such as with CVD, glucose metabolism (glycolysis and the pentose phosphate pathway) typically increases relative to fatty acid oxidation [195–197]. Further longitudinal research exploring multi-omic markers of carbohydrate metabolism in response to TRAP exposure would help clarify the salient relationships.

#### 3.2.3 Amino Acid Metabolism

Although no methylomic or proteomic markers related to the alanine, aspartate, and glutamate metabolism KEGG pathway were identified as significantly associated with TRAP, our MetaboAnalyst pathway analysis synthesizing results from across studies identified the alanine, aspartate, and glutamate metabolism KEGG pathway as significantly enriched by metabolites associated with short and long-term TRAP exposure (p = 3.39 × 10^−4^ and p = 6.0 × 10^−3^, respectively). There were 14 metabolites representing the KEGG pathway, but there were no consistent patterns of associations among short- and long-term TRAP exposure and concentrations of these metabolites [80,94,95,97,105,108,129–131,163,164,167,176,191] (Table 2, Supplementary 2 Tables 6-7).

The arginine and proline metabolism KEGG pathway was represented by biomarkers of all three omics types (two genes, one protein, and 14 metabolites) (Table 2) and our MetaboAnalyst pathway analysis synthesizing the metabolomics literature suggested this pathway was significantly enriched by metabolites significantly associated with short-term TRAP exposure (p = 6.62 × 10^−4^), but not long-term TRAP exposure. Taken together, there is moderately strong evidence that arginine and proline metabolism may affect the relationship between TRAP and CVD. For example, in the methylomics literature, exposure to short-term PM_2.5_ was associated with hypomethylation of the genes that code for nitric oxide synthase 2 (*NOS2*) and arginase 2 (*ARG2*) [58,117,131]. These are key enzymes for macrophage pathways linking L-arginine metabolism to inflammation and immunity [198]. The protein NOS2 catalyzes the reaction of L-arginine to nitric oxide (NO), which inhibits cell proliferation and kills pathogens [199,200]. The protein ARG2 catalyzes the reaction of L-arginine to L-ornithine, which can metabolize further into polyamines and L-proline. Notably, L-ornithine production promotes cell proliferation and repairs tissue damage [201,202]. ARG2 activity is also associated with the killer-type macrophage response [198,203,204]. Many of the metabolites related to this arginine and proline metabolism pathway were implicated across the metabolomics literature – though some of the results were inconsistent in terms of direction of association (Supplementary 2 Table 6) [80,94,95,98,105,108,130,132,163,164,194]. For example, short-term PM_2.5_ was associated with lower levels of L-arginine, L-glutamate, phosphocreatine, and pyruvate, and with higher levels of L-ornithine and nitric oxide [80,94,98,110,146]. However, short-term O_3_ exposure was associated with higher levels of creatinine, L-arginine, L-glutamate, L-ornithine, and L-proline [110,176]. Furthermore, other short-term PM exposures were associated with lower levels of creatinine, and higher levels of L-arginine, L-glutamate, L-ornithine, L-proline, D-proline, and sarcosine [132,191]. Finally, in the proteomics literature, short-term diesel exhaust was associated with lower levels of the protein creatine kinase B-type [126], and in our network analysis for short-term exposure to TRAP, the protein creatine kinase B-type was also associated with a metabolite related to lipid metabolism (Figure 5). Given the overlap in the biomarkers identified using the three omics types, further research is warranted into how TRAP exposure may plausibly result in clinically meaningful biological cascades involving arginine and proline metabolism. Such an undertaking would require repeated measures of exposures and omics markers to ensure that the relevant temporal relationships are captured for different levels of biology along the pathway (e.g., how methylation changes related to *NOS2* and *ARG2* could affect protein expression and subsequent metabolic processes). Future work should also explore the potential connections among amino acid metabolism (blue symbols), coagulation (yellow symbols), inflammation (red symbols), and endothelial pathways (pink symbols) given the results of our network analyses for both short- and long-term TRAP exposures (Figures 3-6).

#### 3.2.4 Inflammation and Immunity

Many methylomic and proteomic markers (but not metabolomic markers) identified in the literature review as associated with TRAP exposure were involved with pathways involved in inflammation and immunity (Figure 2). The most enriched pathways included cytokine and chemokine signaling, toll-like receptor (TLR) signaling, and mitogen-activated protein kinase (MAPK) signaling. Biomarkers of these pathways (especially of the cytokine and chemokine signaling pathway) were also well-represented in our network analyses (Figures 3-6; red symbols correspond to inflammation and immunity).

Our Reactome pathway analysis identified cytokine signaling in the immune system as significantly enriched by genes related to the methylation sites and proteins associated with short-term TRAP exposure (p = 1.11 × 10^−16^ and p = 1.11 × 10^−16^, respectively). This pathway was also significantly enriched by proteins associated with long-term TRAP exposure (p = 1.11 × 10^−16^), but not genes related to the methylation sites. In particular, there were 13 genes and 40 proteins (with 10 overlapping gene-protein markers) that were part of the cytokine-cytokine receptor interaction KEGG pathway, as well as eight genes and 19 proteins (with four overlapping gene-protein markers) that were part of the chemokine signaling KEGG pathway (Table 2). Short-term PM_2.5_ exposure was associated with hypermethylation of the genes encoding for cytokines and chemokines, such as interleukin 6 (*IL6*), interleukin 10 (*IL10*) granulocyte-macrophage colony-stimulating factor 2 (*CSF2*), fractalkine (*CX3CL1*), interferon-gamma inducible protein 10 (*CXCL10*), and macrophage inflammatory protein 1 alpha (*CCL3*) [58,115]. In contrast, short-term PM_2.5_ was associated with hypomethylation of the genes that encode monocyte chemoattractant protein 1 (*CCL2*) and cluster of differentiation 40 ligand (*CD40LG*) [58,115,120,141,142]. Additionally, long-term PM_2.5_ exposure was associated with hypomethylation of tumor necrosis factor (*TNF*) and *TNF* receptor superfamily member 13C (*TNFRSF13C*) [45,141]. Consistent with some but not all of the methylation trends, proteomics studies found that both short- and long-term exposure to TRAP was associated with higher levels of most cytokine and chemokine proteins (exceptions included inverse associations with tumor necrosis factor receptor superfamily member 11B, interleukin 4, interleukin 8, and eotaxin-1) [73,79,81,86,88,90,90,91,113,115,120,124,132,141,162,205–209]. These observations were consistent across pollutants and exposure windows. Additional research on the associations among pollutants other than PM_2.5_ and the methylation of genes encoding for cytokines and chemokines would further strengthen the already compelling evidence that TRAP may impact cytokine and chemokine signaling in ways that could affect respiratory and cardiovascular outcomes. Cytokines and chemokines regulate the immune response by controlling immune cell trafficking and the cellular arrangement of immune organs [210,211]. High levels of both cytokines and chemokines represent immune activation and inflammation, and are predictive of CVD and adverse cardiovascular events, such as heart failure and myocardial infarction [211–214]. In addition, many of the key cytokines identified here are involved in the pathogenesis of asthma, COPD, and pulmonary fibrosis [215]. Finally, as shown in our network analyses, many of the genes and proteins associated with short-term TRAP exposure (e.g., *IL6*/IL6*, CXCL10/*CXCL10*, CCL2*/CCL2) were inter-connected, and were also connected to metabolites of amino acid and lipid metabolism (Figures 5 and 6) – strengthening the argument for involvement of cytokine signaling in the physiological response to TRAP.

Eight methylomic markers and 11 proteomic markers, with four overlapping gene-protein markers and no metabolomic markers, represented the TLR signaling KEGG pathway (Table 2). Short-term exposure to PM_2.5_ and BC were associated with hypomethylation and hypermethylation of *TLR2*, respectively [38,58]. Exposure to short-term PM_10_ and other short-term TRAP was associated with hypomethylation of *TLR4* [144,145]. Exposure to short-term PM_10_ and SO_4_ were associated with hypomethylation of *CD14* and *MAP3K7*, respectively [43,145]. The remaining methylomic and proteomic markers belonging to the TLR KEGG pathway overlapped with the cytokine-cytokine receptor interaction KEGG pathway described previously and in Table 2. These trends are important because the TLR signaling pathway detects pathogen-associated molecular patterns, stimulating both the nuclear factor kappa-light-chain-enhancer of activated B cells (NF-kB) and MAPK pathways, as well as cytokine production, thereby affecting inflammatory and immune responses associated with CVD and adverse respiratory outcomes [216,217].

In addition to the trends for cytokine and chemokine signaling and the TLR signaling pathways, we identified 12 methylomic markers and nine proteomic markers associated with TRAP as belonging to the MAPK signaling KEGG pathway, with two overlapping gene-protein markers and no metabolomic markers (Table 2). In the methylomics literature, short-term BC exposure was associated with hypermethylation of *MAP3K2* and *MAP3K6*, as well as hypomethylation of *MAP4K3* and *MKNK2* [43]. Short-term SO_4_ exposure as associated with hypermethylation of *MAP3K11*, and hypomethylation of *RPS6KA3*, *MAP3K7*, and *TGFB1* [43]. Long-term exposure to PM_10_ and NO_2_ were associated with hypomethylation and hypermethylation of *PDGFB* and *CACNA2D1*, respectively [45,53]. Lastly for the methylomics literature, short-term PM_2.5_ exposure was associated with hypermethylation of *FGF2* [115]. In the proteomics literature, short-term PM_2.5_, UFP, BC, NO_2_, and CO exposures were associated with higher levels of fibroblast growth factor 2 protein [115,132]. In addition, short-term diesel exhaust exposure was associated with higher levels of MAPK 1 and cell division control protein homolog 42, and lower levels of protein kinase C beta type [126,162]. Finally, short-term UFP, BC, NO_2,_ and CO were associated with higher levels of tropomyosin receptor kinase B [132]. Synthesizing across the studies, our Reactome pathway analysis identified the MAPK signaling cascades pathway as significantly enriched by proteins associated with short-term TRAP exposure (p = 4.35 × 10^−8^). Although this pathway was not significantly enriched by methylation markers associated with TRAP exposure, the body of evidence taken together suggests that TRAP exposures may affect MAPK signaling cascades – which is critical since this pathway has implications for oxidative stress, vascular remodeling and dysfunction, cardiac hypertrophy, cardiac remodeling, and atherosclerosis [218–223].

#### 3.2.5 Coagulation

The complement and coagulation cascades KEGG pathway was represented by four methylomic markers and 14 proteomic markers significantly associated with TRAP, with two overlapping gene-protein markers. There were no metabolomic markers of this pathway identified as significantly associated with TRAP (Table 2). Short-term exposure to PM_2.5_ was associated with hypomethylation of the genes that encode plasminogen activator inhibitor type I (*SERPINE1*), coagulation factor III (*F3*), and coagulation factor II receptor (*F2R*), as well as hypermethylation of coagulation factor II (*F2*) [38,46,120,142,157]. Within the proteomics literature, short-term exposure to PM_10_ and PM_2.5-10_ were associated with lower levels of the protein plasminogen activator inhibitor type 1, whereas long-term exposure to PM_2.5_, NO_2_, CO, and O_3_ were associated with higher levels of this protein [69,71,73]. Additionally, short-term exposure to PM_2.5_, UFP, BC, NO_2_, and CO were associated with higher levels of coagulation factor III protein (F3) [123,132]. The combination of associations with short-term exposures and methylation markers and long-term exposures and proteins (e.g., *SERPINE1*) may provide evidence for time-dependent biological cascades or effects; future research should explore this possibility using a study design that can take advantage of repeated measures for exposures and outcomes. Further research could explore the possibility of similar overlap across omics types by building on the TRAP and proteomics literature suggesting significant and generally positive associations with other key coagulation and complement proteins (e.g., complement component 3, complement component 4B, fibrinogen, Von Willebrand factor, coagulation factor VII, D-dimer, alpha-1 antitrypsin, protein C inhibitor, complement C1q subcomponent subunit A, and tissue-type plasminogen activator; Supplementary 2 Table 5) [70,73,75,83,89,119–122,126,132,209,224,225]. The importance of complement and coagulation cascades are also underscored by the connections of coagulation factors, coagulation factor responses, plasminogen activators, and plasminogen activator inhibitors in the network analyses (represented by yellow markers) to biomarkers of amino acid metabolism (blue markers), lipid metabolism (green markers), and inflammation and immunity (red markers; Figures 3-6). Taken together, there is strong evidence supporting the putative links between TRAP exposure, coagulation and complement cascades, and CVD (Figure 2). This is supported by other studies that show that higher levels of plasminogen activator inhibitor 1, fibrinogen, Von Willebrand factor, coagulation factor VII, and complement component 3 are each associated with risk of CVD and atherosclerosis [222,223,226–231]. Furthermore, higher levels of plasminogen activator inhibitor 1 and Von Willebrand factor have been associated with increased odds of myocardial infarction [222,231].

#### 3.2.6 Endothelial Function

Methylomic, proteomic, and metabolomic markers associated with TRAP exposure were associated with five KEGG pathways related to endothelial function: cell adhesion molecules, vascular endothelial growth factor (VEGF) signaling, vascular smooth muscle contraction, lipid and atherosclerosis, and leukocyte transendothelial migration (Table 2).

The first KEGG pathway, cell adhesion molecules, was represented by five methylomic markers, five proteomic markers (including three overlapping with the methylomic markers), and no metabolomic markers (Supplementary S2 Tables 4 and 5). The three overlapping markers were cluster of differentiation 40 ligand (CD40LG), p-selectin (SELP), and intercellular adhesion molecule 1 (ICAM1). For CD40LG, short-term PM_2.5_ was associated with hypomethylation of the corresponding gene [115,120,142], whereas short-term PM_2.5_, NO_2_, SO_2_, SO_4_, EC, and multiple PM components were associated with higher levels of the protein [73,113,113,115,120,141,142,206,209,225]. For SELP, long-term PM_2.5_ was associated with hypomethylation of the corresponding gene and long-term PAHs were associated with lower levels of the protein [45,120,209,232]. For ICAM1, short-term BC and O_3_ were associated with hypomethylation of the corresponding gene [38], short-term PM_2.5_ had inconsistent associations with the corresponding gene [38,58,120,141], and both short- and long-term TRAP exposures were generally associated with higher levels of the protein [89,121,141,206,207,233–236]. Biomarkers of the cell adhesion molecule pathway (e.g., SELP, ICAM1) were also identified in our network analysis for both short- and long-term TRAP exposures as being highly connected to markers of other biological processes (e.g., lipid metabolism; Figures 3-6). Cell adhesion molecules are essential in the normal development of the heart and blood vessels; however, they play a role in the development of respiratory and cardiovascular diseases such as pulmonary fibrosis and atherosclerosis [237].

The second KEGG pathway, the VEGF signaling pathway, was represented by no methylomic, three proteomic, and two metabolomic markers associated with TRAP exposure (Supplementary S2 Tables 5 and 6). For proteomics, short-term exposure to diesel exhaust was associated with higher levels of the cell division control protein 42 homolog, and lower levels of protein kinase C beta type [126]. In addition, exposure to short term-term NO_2_ and long-term NO_x_ were associated with higher levels of VEGF-alpha (VEGFA) [81,113]; VEGFA was also identified as connected to markers of lipid metabolism and amino acid metabolism in our network analysis for short-term TRAP exposure (Figure 5). For metabolomics, short-term PM_2.5_ was associated with higher levels of nitric oxide, and short-term EC was associated with higher levels of prostaglandin I2 [117,146,164]. Up-regulation of VEGF signaling is involved in angiogenesis and can be a response to hypoxia [238]. Higher concentrations of these analytes associated with TRAP exposure could indicate difficulty delivering oxygen from the lungs to the periphery; however, VEGF signaling is not always pathological.

The third KEGG pathway, vascular smooth muscle contraction, was represented by one methylomic, three proteomic, and four metabolomic markers associated with TRAP exposure (Supplementary S2 Tables 4-6). For methylomics, long-term PM_2.5_ was associated with hypomethylation of guanine-nucleotide binding protein, alpha subunit complex locus (*GNAS*) [45]. For proteomics, short-term UFP, BC, NO_2_, and CO were associated with higher levels of endoglin [132], and short-term diesel exhaust was positively associated with mitogen activated protein kinase 1 and negatively associated with protein kinase C beta type [126]. For metabolomics, short-term PM_2.5_ was positively associated with nitric oxide and 20-hydroxyeicosatetraenoic (HETE) acid [107,117,146], and short-term TRAP was positively associated with arachidonate and prostaglandin I2 [164,165,170]. Contraction of the vascular smooth muscle within arteries, arterioles, veins, and lymphatic vessels increases resistance in the cardiovascular system, and decreases blood flow [239]. TRAP-associated modulation in these signals could inform part of the relationship between TRAP exposure and blood pressure, and therefore CVD. Further research is needed to clarify the exact physiological mechanisms linking TRAP, omics signals, blood pressure, and CVD.

The fourth KEGG pathway, lipid and atherosclerosis, was represented by no methylomic or proteomic markers but three metabolomic markers associated with TRAP exposure (Supplementary S2 Table 6). Short-term PM_2.5_ was positively associated with nitric oxide, and short-term TRAP was positively associated with cholesterol and triglyceride [117,132,146]. Cholesterol and triglycerides, both positively associated with TRAP exposure, are risk factors for atherosclerosis. Furthermore, TRAP is already known to be associated with atherosclerosis through exacerbation of risk factors such as hypertension and insulin resistance [240].

The final KEGG pathway, leukocyte transendothelial migration, was represented by three methylomic markers, six proteomic markers (one overlapping with a methylomic marker), and no metabolomic markers associated with TRAP exposure (Supplementary S2 Tables 4 and 5). The trends for the overlapping marker (ICAM1), as well as two of the other proteomic markers (i.e., protein kinase C beta type and cell division control protein homolog 42) were described previously. The other methylation markers associated with short-term PM_2.5_ encode for protein subunit alpha 13 (positive association) and actinin alpha 3 (negative association) [40,158]. The other proteomic markers positively associated with short-term TRAP exposure included vascular cellular adhesion molecule 1 (VCAM1; with PM_2.5_, NO_2,_ CO, SO_4_, and O_3_) [89,121,206], matrix metalloproteinase (MMP2; with BC and PNC), and MMP9 (with SO_2_) [79]. In our network analysis for short-term TRAP exposures, MMPs shared network connections with markers of processes such as lipid and amino acid metabolism (Figure 5). Leukocyte trans-endothelial migration is critical in the immune response and responsible for a facilitating a systemic reaction upon exposure to a pathogen [241]. The subclinical effects of differential leukocyte count post TRAP exposure have previously been noted [242], and represent part of the well-documented inflammatory response to TRAP.

#### 3.2.7 Oxidative Stress

Multiple KEGG pathways represented in the methylomic, proteomic, and metabolomic literature are associated with the oxidative stress response (Table 2; Figure 2). For example, the citrate cycle, pentose phosphate metabolism, MAPK signaling, p53 signaling, Janus Kinase/signal transducers and activators of transcription (JAK-STAT) signaling, apoptosis, and regulation of autophagy KEGG pathways are all known to be activated in response to oxidative stress [219,243–249]. The biomarkers related to several of these pathways were described previously. Others are described in this section.

The p53 signaling pathway is activated in response to oxidative stress and TRAP exposure, and helps to ensure cell survival [244,245]. For this pathway, one methylomic and seven proteomic markers (including one overlapping gene-protein marker) were identified as significantly associated with TRAP (Supplementary 2 Tables 4, 6, 7). Short-term exposure to PM_2.5_ was associated with hypomethylation of *SERPINE1* [142]. Additionally, short-term exposure to PM_10_ and PM_2.5-10_ was associated with lower levels of the corresponding protein, whereas long-term exposure to PM_2.5_, PM_2.5-10_, NO_2_, CO, and O_3_ were associated with higher levels [69,71,73]. Furthermore, short-term BC and NO_2_ were associated with higher levels of insulin-like growth factor binding protein 1 and 3, while short-term diesel exhaust was associated with lower levels of insulin-like growth factor binding protein 2 and 14-3-3 protein sigma [79,126]. Finally, long-term PM_2.5_ and PM_10_ exposures were associated with higher levels of alpha-1 antitrypsin [83]. Given the role of p53 signaling in anti-angiogenesis, programmed cell death, metabolism regulation, and vasodilation, this pathway can affect cardiovascular outcomes [250,251]. In addition, p53 signaling plays a supportive role in the maintenance of lung homeostasis; therefore, dysregulation and deficiency of p53 signaling can be associated with respiratory diseases [252].

Similarly to the p53 signaling pathway, the JAK-STAT signaling pathway is activated by oxidative stress and reactive oxygen species [248]. This signaling pathway is mainly involved in coordinating immune responses, including cytokine signaling [253]. Four methylomic markers and fourteen proteomic markers (including four overlapping gene-protein markers) of this pathway were identified as significantly associated with TRAP (Supplementary 2 Tables 4, 6, 7). Three of the methylomic markers (for genes *CSF2, IL6,* and *IL10*) were described in the section on inflammation and immunity. Briefly, short- and long-term TRAP was associated with hypomethylation of these markers and higher levels of the proteins they encode [58,73,88–91,113,115,124,141,162,205,225,235,254,255]. Hypermethylation of one methylomic marker relevant here (related to a gene that encodes interferon gamma (*IFNG*)) was associated with short-term TRAP exposure (though short-term BC was associated with hypomethylation) [38,256]. Relatedly, short-term PM_2.5_, NO_2_, CO, PAHs, and PM constituents were associated with higher levels of the protein IFNG [113,205]. Short-term TRAP was also positively associated with other proteins involved in JAK-STAT signaling including granulocyte colony-stimulating factor 3, granulocyte-macrophage colony-stimulating factor receptor alpha, interleukin 2 alpha, interleukin 5, interleukin 7, interleukin 12, and signal transducer and activator of transcription 3 (STAT3) [113,115,124,132,162,208]. In contrast, short-term TRAP was associated with lower levels of interleukin 4, interleukin 13, and protein tyrosine phosphatase non-receptor type 6 [113,115,162]. These associations with markers related to JAK-STAT signaling are important for the relationship between TRAP exposure and CVD outcomes because dysregulation of JAK-STAT signaling is associated with CVD [257,258]. Furthermore, cytokine signaling induced through the JAK-STAT pathway is implicated in COPD, asthma, and other respiratory conditions [259,260].

Apoptosis, or programmed cell death, can be caused by oxidative stress [246]. Representing the apoptosis KEGG pathway, TRAP was associated with one methylomic marker, three proteomic markers (including one overlapping with a methylomic marker), and one metabolomic marker (Supplementary 2 Tables 4-6). Trends for the overlapping methylomic-proteomic marker, tumor necrosis factor alpha, were described previously. For the other proteomic markers, short-term PM_10_, UFP, NO_2_, CO, and PAHs were positively associated with interleukin 1 beta, whereas short-term UFP, BC, NO_2_, and CO were inversely associated with tropomyosin receptor kinase B [91,132,205]. For metabolomics, short-term PM_2.5_ and UFP and long-term PM_2.5_ were associated with lower levels of the sole metabolite, sphingosine [95,98,168]. Apoptosis is a vital part of normal cell turnover and immune system functioning, implicating this pathway in cardiorespiratory disease [261–263].

The final oxidative-stress related KEGG pathway, the regulation of autophagy, is involved in apoptosis and helps maintain cellular homeostasis [246,247,249,264]. This pathway was represented by one methylomic marker and two proteomic markers (including one overlapping marker) associated with TRAP. Trends were previously described for the overlapping marker, interferon-gamma. The other protein, interferon alpha 2, was positively associated with short-term PM_2.5_ [115]. Proper functioning of adaptive autophagy processes is important for cardiovascular health and aging [265–267].

#### 3.2.8 TRAP, Omics, and Respiratory Disease

Short- and long-term TRAP exposure is associated with worse respiratory outcomes, including worse lung function [58,87,108,151,268–272], and with more asthma exacerbation and COPD burden [271,273–276]. In our review, three methylomic markers, seven proteomic markers (including three overlapping methylomic-proteomic markers), and three metabolomic markers were represented in the KEGG pathway for asthma (Table 2). The overlapping markers included three inflammation and immunity markers (TNF, CD40LG, and IL-10); we described trends for these previously [58,69,88,91,113,115,120,124,141,205,206,206,209,235,255,269,277]. For the other proteomics markers, short-term PM_2.5_, PM_10_, NO_2_, CO, and SO_2_ were inversely associated with IL-4, short-term CO was inversely associated with IL-13 [113,124,278]; and short-term NO_2_ and diesel exhaust were positively associated with IL-5 [113,124]. Additionally, short-term PM_2.5_, PM_10_, CO, and SO_2_ were inversely associated with monocyte chemoattractant protein 1 whereas long-term PM_2.5_, NO_2_, and NO_x_ were positively associated with this protein [81,113,115]. For metabolomic markers, short-term TRAP was positively associated with leukotriene C4, and inversely associated with prostaglandin D2 [165]; and short-term NO_2_, CO, and EC were inversely associated with histamine [163]. These trends, along with others described in previous sections suggest plausible biological processes that affect the TRAP exposure-respiratory disease relationship. For example, it has been observed that linoleate metabolism is associated with asthma [101], and arginine and proline metabolism as well as methionine and cysteine metabolism are associated with asthma and COPD [103]; these are processes associated with TRAP exposures. Additionally, elevated NO is characteristic of airway inflammation [279] and we previously described trends relating TRAP to higher NO [58,117,146]. Similar trends are observed between TRAP exposures and markers of systemic inflammation (e.g., CRP, fibrinogen) that are associated with worse lung function [280–283]. Finally, the associations we described previously relating TRAP exposures to cytokines and chemokines have implications for airway remodeling asthma, and COPD [215].

#### 3.2.9 TRAP, Omics, and CVD

As described above and elsewhere, many studies have observed associations between TRAP exposure and biomarkers related to CVD (e.g., [284–286]). A subset of studies used meet-in-the-middle approaches (i.e., identifying common associations of exposures and CVD outcomes with biomarkers), mediation analyses, and other approaches to more directly link TRAP exposures to CVD outcomes (e.g., heart rate [256], blood pressure [143,144], incident CVD [81]). As in our review, these studies considered biomarkers for processes related to inflammation and immunity, endothelial function, and oxidative stress. Most of these studies considered only single omic types, but one that considered both proteomic and metabolomic biomarkers identified 20 biomarkers associated with both short-term TRAP and changes in blood pressure or heart rate variability [132]. As in our review, that study identified biomarkers implicated in lipid metabolism (e.g., trimethylamine N-oxide), cellular energy production (e.g., succinic acid), inflammation (e.g., C-reactive protein), coagulation (tissue factor pathway inhibitor), endothelial function (e.g., angiotensin-converting enzyme), and oxidative stress (e.g., malondialdehyde). Our review was able to take this type of logic one step further—with the network analyses (Figures 3-6). By integrating information across multi-omic types, we can build on the systems biology approaches now being used to understand the pathophysiology of CVD (e.g., [287,288]). Specifically, our network analyses suggest that interconnections among amino acid metabolism, lipid metabolism, inflammation, coagulation, and endothelial function are important to the relationship between TRAP exposures and CVD.

## 4. Conclusions

To our knowledge, this is the first systematic review synthesizing the literature focused on TRAP-associated methylomic, proteomic, and metabolomic biomarkers in the context of respiratory and cardiovascular outcomes. Through a comprehensive, integrated lens, we explored TRAP-associated pathways involving lipid metabolism, cellular energy production, amino acid metabolism, inflammation and immunity, coagulation, endothelial function, and oxidative stress. We find that a multi-omics synthesis provides new insights into the biological pathways associated with TRAP and has advantages over single-omic approaches. Synthesizing results from the (predominately single-omic) literature, we showed that similar or analogous biomarker signals were observed across multiple omic types (e.g., TRAP exposure associated with methylation of genes encoding for proteins that are also associated with TRAP). Specifically, we identified consistent patterns between methylation status and protein levels within cytokine-cytokine signaling, TLR signaling, MAPK signaling, complement and coagulation cascades, cell adhesion molecules, and asthma KEGG pathways. Additionally, we observed analogous proteomic and metabolomic associations with TRAP exposure within certain lipid and amino acid metabolism KEGG pathways. Finally, within the arginine and proline metabolism KEGG pathway, we were able to integrate methylomic, proteomic, and metabolomic findings together to provide evidence suggesting possible mechanistic linkages between TRAP exposure, subclinical indicators, and clinical disease. Corroborating evidence across multiple levels of biology – including with a focus on functional interrelationships and network analyses – is only possible with multi-omics. Furthermore, multi-omics has the potential to aid in the discovery and assessment of quantitative biomarkers at different levels of biology (related methylation patterns, proteins, and metabolites) that could predict subclinical and perhaps clinical respiratory and cardiovascular responses to TRAP exposure, thereby improving clinical and public health decision making. This could perhaps be clinically translated using advances to epigenetic clocks and other risk prediction tools.

### 4.1 Strengths and Limitations

A major strength of our systematic review is that we provided a synthesis of findings from across three types of omics markers. This multi-omics process offers superior insight into the biological underpinnings of respiratory and cardiovascular disease than single-omics methods alone. We compiled methylomic, proteomic, and metabolomic evidence from methodologically diverse studies in a novel way to understand how short- and long-term TRAP exposure-associated multi-omics signals relate to one another, allowing us to identify the most relevant biological pathways that may be involved in the pathogenesis of cardiorespiratory disease. Nevertheless, our review had several limitations. First, to synthesize results across studies that used heterogeneous exposure metrics and methods, we made the simplifying assumption of categorizing short- and long-term exposures as ≤30 days and >30 days, respectively. This decision was supported by convention within the literature but does not necessarily reflect a critical biological change occurring at 30 days. Additionally, due to the availability of published studies, there were fewer long-term exposures represented in our analysis. Second, to synthesize the biological implications of the individual biomarkers identified as associated with TRAP, we made simplifying assumptions that we could include all individually identified biomarkers together in our pathway and network analyses, and although we considered short- and long-term exposures separately, we did not separate results by pollutant type. It is likely that different TRAP components have different biological impacts. More generally, it is possible that direct comparisons or synthesis were not warranted in each case due to certain differences in study population, exposure metric, or other methodological choices within the individual studies that would result in meaningful differences in the true underlying biology. Third, our synthesis of the results and identification of relevant pathways was necessarily limited by the choices of the individual studies. If the studies did not include certain biomarkers that may in fact be important to the physiological response to TRAP, we could not capture them – particularly for proteomics, this may have limited our findings since there were somewhat fewer studies with large numbers of proteins assayed and the literature may have overrepresented certain biological pathways due to precedent rather than biology. Future studies may consider a more comprehensive set of proteins. Similarly, if metabolite identification with a high level of confidence was not provided by the individual studies, we may have missed critical biological pathways. Third, we limited the scope of our review to exclude people who were pregnant and/or under 18 years old. Future research should consider these important populations. Finally, and perhaps most critically, we could not assess whether TRAP exposures resulted in meaningful biological cascades following the gene to protein to metabolite paradigm as no study we reviewed included all three omics types and none included the repeated measures of the omics markers that would be needed to assess dynamic biological processes.

### 4.2 Future Directions

Building on the strengths of the studies presented in this review and the conclusions that could be drawn by comparing the results using heterogenous research methodologies, several critical areas for further research are warranted. The primary challenges our field currently faces are related to the true integration of multi-omics signals within studies that can appropriately characterize the dynamic and complex biological processes linking TRAP exposure to subclinical and clinical disease. To address this critical challenge, we need large, longitudinal studies representing diverse study populations. Ideal features include time-varying, high-resolution exposure assessment coupled with repeated quantification of multi-omics signals in multiple tissue types with comprehensive assay coverage. If multiple cohorts are included in a study, standardization of methods across cohorts would facilitate interpretation and comparability of results. A major goal of such a study would be to consider how air pollution exposures might lead to physiological signals suggestive of the biological cascades leading from exposure to subclinical disease to clinical disease. A consideration of both the short- and long-term physiological effects of TRAP would be warranted (including a consideration of individual TRAP components and TRAP mixtures). It would also be worth examining sex and gender differences, along with other differences that could lead to disparities in health consequences attributed to air pollution exposure. The use of emerging and novel data management and analysis approaches that can handle large and complex data structures inherent in multi-omics studies will be important (e.g., multiblock methods and tensor decomposition methods) [24,287,289–294]. Finally, it would be critical to consider the optimal multi-omics integration approach (e.g., whether each omics type is analyzed first and then types are synthesized, or whether processing integrates across omics types earlier) [295–297]. If such a comprehensive study could be conducted, it would provide mechanistic insight into the pathophysiology and progression of disease and would inform identification of multi-omic signatures of air pollution exposure that could be predictive of key health outcomes. Insights gained from such studies could inform screening priorities, clinical decision-making, and public policy.

## Author Contributions

Conceptualization, F.K., C.U. and L.C.; methodology, C.C., F.K., K.K., and L.C.; formal analysis, C.C.; investigation, C.C., F.K. and C.U.; resources, L.C.; data curation, C.C., F.K., and C.U.; writing— original draft preparation, C.C., F.K., K.K., and L.C..; writing—review and editing, all authors; visualization, C.C., F.K, and C.U,; supervision, L.C.; funding acquisition, L.C. All authors have read and agreed to the published version of the manuscript.

## Institutional Review Board Statement

Not applicable

## Data Availability Statement

The datasets generated and analyzed for this study can be found in the manuscript and supplements.

