## Supplementary S1 for "Methylomic, proteomic, and metabolomic correlates of traffic-related air pollution: A systematic review, pathway analysis, and network analysis relating traffic-related air pollution to subclinical and clinical cardiorespiratory outcomes"

**Methylation Search 1**: EMBASE Search Strategy: English Language Articles published between 2010-2023. Search conducted on 01/11/23

#1 ('methylation'/mj/exp OR 'methylate' OR dna) AND 'methylation'/mj/exp OR 'methylation' OR 'epigenetics'/mj/exp OR 'epigenetics' OR methylation OR 'methylome'/mj/exp OR 'methylome'

#2 'air pollution'/mj/exp OR 'air pollution' OR 'traffic related air pollution'/mj/exp OR 'traffic related air pollution' OR 'air pollutant'/mj/exp OR 'air pollutant' OR 'traffic pollution'/mj/exp OR 'traffic pollution'

#3 #1 AND #2

#4 #3 AND (2010:py OR 2011:py OR 2012:py OR 2013:py OR 2014:py OR 2015:py OR 2016:py OR 2017:py OR 2018:py OR 2019:py OR 2020:py OR 2021:py OR 2022:py) AND ('case control study'/de OR 'clinical article'/de OR 'cohort analysis'/de OR 'comparative study'/de OR 'controlled study'/de OR 'correlational study'/de OR 'cross sectional study'/de OR 'crossover procedure'/de OR 'experimental study'/de OR 'human experiment'/de OR 'longitudinal study'/de OR 'major clinical study'/de OR 'multicenter study'/de OR 'observational study'/de OR 'panel study'/de OR 'prospective study'/de OR 'questionnaire'/de OR 'randomized controlled trial'/de OR 'retrospective study'/de) AND ([adult]/lim OR [aged]/lim OR [middle aged]/lim OR [very elderly]/lim OR [young adult]/lim) AND [english]/lim AND ([adult]/lim OR [young adult]/lim OR [middle aged]/lim OR [aged]/lim OR [very elderly]/lim) AND [humans]/lim

**Results in 219 publications**

**Methylation Search 2**: PubMed Search Strategy: English Language Articles published between 2010-2023. Search conducted on 01/11/23

(("methyl*"[Title]) AND ("humans"[MeSH Terms] AND "english"[Language] AND "adult"[MeSH Terms]) AND (("pollution"[Title] OR "traffic related air pollution"[Title] OR "traffic pollution"[Title] OR "TRAP"[Title] OR "pollutants"[Title] OR "air pollutants"[Title] OR "air pollution"[Title] OR "particulate matter"[MeSH Terms] OR "air pollutants"[MeSH Terms]) AND ("humans"[MeSH Terms] AND "english"[Language] AND "adult"[MeSH Terms]))) AND (humans[Filter])

**Results in 138 publications**

**Proteomics Search 1:** EMBASE Search Strategy: English Language Articles published between 2010-2023. Search conducted on 11/30/22

#19 AND (2010:py OR 2011:py OR 2012:py OR 2013:py OR 2014:py OR 2015:py OR 2016:py OR 2017:py OR 2018:py OR 2019:py OR 2020:py OR 2021:py OR 2022:py) AND ('case control study'/de OR 'clinical article'/de OR 'cohort analysis'/de OR 'comparative study'/de OR 'controlled study'/de OR 'correlational study'/de OR 'cross sectional study'/de OR 'crossover procedure'/de OR 'experimental study'/de OR 'human experiment'/de OR 'longitudinal study'/de OR 'major clinical study'/de OR 'multicenter study'/de OR 'observational study'/de OR 'panel study'/de OR 'prospective study'/de OR 'questionnaire'/de OR 'randomized controlled trial'/de OR 'retrospective study'/de) AND ([adult]/lim OR [aged]/lim OR [middle aged]/lim OR [very elderly]/lim OR [young adult]/lim) AND [english]/lim

#19 = #16 AND #18

#16 = 'proteins'/mj OR 'protein'/mj OR 'proteomics'/mj OR proteomic OR 'proteome'/mj OR 'inflammation'/mj

#18= 'air pollution'/mj OR 'traffic related air pollution'/mj OR 'air pollutant'/mj OR 'traffic pollution'/mj

**Results in 160 publications**

**Proteomics Search 2:** PubMed Search Strategy: English Language Articles published between 2010-2023. Search conducted on 12/06/22

(("prote*"[Title]) AND ("humans"[MeSH Terms] AND "english"[Language] AND "adult"[MeSH Terms]) AND (("pollution"[Title] OR "traffic related air pollution"[Title] OR "traffic pollution"[Title] OR "TRAP"[Title] OR "pollutants"[Title] OR "air pollutants"[Title] OR "air pollution"[Title] OR "particulate matter"[MeSH Terms] OR "air pollutants"[MeSH Terms]) AND ("humans"[MeSH Terms] AND "english"[Language] AND "adult"[MeSH Terms]))) AND (humans[Filter])

**Results in 166 publications**

**Metabolomics Search 1:** EMBASE Search Strategy: English Language Articles published between 2010-2023. Search conducted on 12/15/22

#1 'metabolites'/mj/exp OR 'metabolites' OR 'metabolite'/mj/exp OR 'metabolite' OR 'metabolomics'/mj/exp OR 'metabolomics' OR metabolomic OR 'metabolome'/mj/exp OR 'metabolome'

#2 'air pollution'/mj/exp OR 'air pollution' OR 'traffic related air pollution'/mj/exp OR 'traffic related air pollution' OR 'air pollutant'/mj/exp OR 'air pollutant' OR 'traffic pollution'/mj/exp OR 'traffic pollution'

#3 #1 AND #2

#4 #3 AND (2010:py OR 2011:py OR 2012:py OR 2013:py OR 2014:py OR 2015:py OR 2016:py OR 2017:py OR 2018:py OR 2019:py OR 2020:py OR 2021:py OR 2022:py) AND ('case control study'/de OR 'clinical article'/de OR 'cohort analysis'/de OR 'comparative study'/de OR 'controlled study'/de OR 'correlational study'/de OR 'cross sectional study'/de OR 'crossover procedure'/de OR 'experimental study'/de OR 'human experiment'/de OR 'longitudinal study'/de OR 'major clinical study'/de OR 'multicenter study'/de OR 'observational study'/de OR 'panel study'/de OR 'prospective study'/de OR 'questionnaire'/de OR 'randomized controlled trial'/de OR 'retrospective study'/de) AND ([adult]/lim OR [aged]/lim OR [middle aged]/lim OR [very elderly]/lim OR [young adult]/lim) AND [english]/lim

**Results in 282 publications**

**Metabolomics Search 2**: PubMed Search Strategy: English Language Articles published between 2010-2023. Search conducted on 12/15/22

(("metabol*"[Title]) AND ("humans"[MeSH Terms] AND "english"[Language] AND "adult"[MeSH Terms]) AND (("pollution"[Title] OR "traffic related air pollution"[Title] OR "traffic pollution"[Title] OR "TRAP"[Title] OR "pollutants"[Title] OR "air pollutants"[Title] OR "air pollution"[Title] OR "particulate matter"[MeSH Terms] OR "air pollutants"[MeSH Terms]) AND ("humans"[MeSH Terms] AND "english"[Language] AND "adult"[MeSH Terms]))) AND (humans[Filter])

**Results in 187 publications**
